# Feasibility of dual phase 99mTc-MDP SPECT/CT imaging in rheumatoid arthritis evaluation

**DOI:** 10.1101/2020.06.10.20126961

**Authors:** Yasser G. Abdelhafez, Felipe Godinez, Kanika Sood, Rosalie J. Hagge, Robert D. Boutin, Siba P. Raychaudhuri, Ramsey D. Badawi, Abhijit J. Chaudhari

## Abstract

**Objectives:** To prospectively demonstrate the feasibility of performing dual-phase SPECT/CT for the assessment of the small joints of the hands of rheumatoid arthritis (RA) patients, and to evaluate the reliability of the quantitative and qualitative measures derived from the resulting images.

**Methods:** A SPECT/CT imaging protocol was developed in this pilot study to scan both hands simultaneously in RA patients, in two phases of 99mTc-MDP radiotracer uptake; namely the soft-tissue blood pool phase (within 15 minutes after radiotracer injection) and osseous phase (after 3 hours). Joints were evaluated qualitatively (normal vs. abnormal uptake) and quantitatively (by measuring the maximum corrected count ratio [MCCR]). Qualitative and quantitative evaluations were repeated to assess reliability.

**Results:** Four participants completed seven studies (all four were imaged at baseline, and three of them at follow-up after 1-month of arthritis therapy). A total of 280 joints (20 per hand) were evaluated. The MCCR from soft-tissue phase scans was significantly higher for clinically abnormal joints compared to clinically normal ones; *p*<0.001, however the MCCR from the osseous phase scans were not different between the two groups. Intraclass Correlation Coefficient (ICC) for MCCR was excellent (0.9789, 95% confidence interval [CI]: 0.9734-0.9833). Intra-observer agreement for qualitative SPECT findings was good for both the soft-tissue phase (kappa=0.78, 95%CI: 0.72-0.83) and osseous-phase (kappa=0.70, 95%CI: 0.64-0.76) scans.

**Conclusion:** Extracting reliable quantitative and qualitative measures from dual-phase 99mTc-MDP SPECT/CT hand scans is feasible in RA patients. SPECT/CT may provide a unique means for assessing both synovitis and osseous involvement in RA joints using the same radiotracer injection.

## Introduction

Synovitis is the hallmark of Rheumatoid Arthritis (RA), and is typically characterized by leukocyte infiltration, hypervascularity, neoangiogenesis, synoviocyte proliferation and fibroblast activation (1). The inflamed synovial membrane may initiate and promote further invasion of cartilage and bone (2). Additionally, synovitis may stimulate osteoclastic differentiation with subsequent cortical bone resorption and breach of the synovium/bone marrow barrier (3, 4). This has been designated as the outside-in hypothesis, in contrast to the inside-out hypothesis, which postulates that joint inflammation originates in the bone marrow (5). The changes in the status of synovial vascularity and bone metabolism are known to typically precede anatomical changes; therefore, non-invasive imaging tools capable of quantifying these processes may offer unique opportunity for RA disease activity evaluation and risk stratification (6).

Skeletal scintigraphy using diphosphonate radiotracers has been used for assessing inflammatory arthritis (7, 8). Blood pool (BP) images acquired shortly after tracer injection reflects local blood flow and soft tissue vascularity, which typically are increased during inflammation (9); while the delayed osseous phase (3-4 h after injection) is reflective of osteoblastic response (10).

Most of the published studies to date employing scintigraphy in RA patients used a single delayed osseous phase scan, in planar, two-dimensional acquisition mode (11, 12). Only a few studies have utilized scanning at multiple timepoints (13) or used three-dimensional acquisition techniques (14-16). The acquisition of dual-phase SPECT/CT could provide insight into RA disease activity in the small joints. However, there is concern that the spatial and contrast resolution afforded by SPECT/CT may not be adequate for scanning of the small joints, such as those of the wrist and hand, that are affected earlier in the RA disease process.

The purpose of this pilot study was to prospectively demonstrate the feasibility of performing dual-phase SPECT/CT for the assessment of the small joints of the hands of rheumatoid arthritis (RA) patients, and to evaluate the reliability of the quantitative and qualitative measures derived from the resulting images.

## Materials and methods

### Study participants

This pilot study was approved by the Institutional Review Board. Written informed consent was obtained from all study participants. The study recruited participants with established rheumatoid arthritis (RA) based on the American College of Rheumatology (ACR) 2010 criteria (17), and who were candidates for starting treatment with first-line disease modifying anti-rheumatic (DMARDs) drugs (N=3) or tumor necrosis factor-alpha (TNF-**α**) blockers (N=1). SPECT/CT scans were conducted at baseline before starting treatment, and again at one month after therapy. Scan timing during the day, waiting interval, positioning and hydration status were matched between the two timepoints. Drug response was assessed by a board-certified rheumatologist using the ACR 20 criteria (17). Additionally, disease activity score for 28 joints (DAS-28) (18) was recorded after 6 months from baseline scans.

### SPECT/CT imaging protocol development

Our first step was to develop means to standardize the position of participant’s body on the scanner bed such that the hands can extend to fall within the field of view (FOV) of gamma cameras; to minimize photon attenuation and maximize coverage. To achieve this task, we designed an immobilization device (Fig. 1) made of 2 low-attenuation detachable hand-shaped thermoplastic molds mounted to acrylic plates using low-density plastic screws. The top edges of the two plates meet at the midline and the front edges converge distally to match the natural orientation of the outstretched hands. The detachable plates are secured by docking them via grooves on a wooden base, that hangs off the edge of the bed, such that it runs for a length of 120 cm under the bed cushion. The patient weight provides stability to the assembly (Fig. 1).

**Fig. 1.**
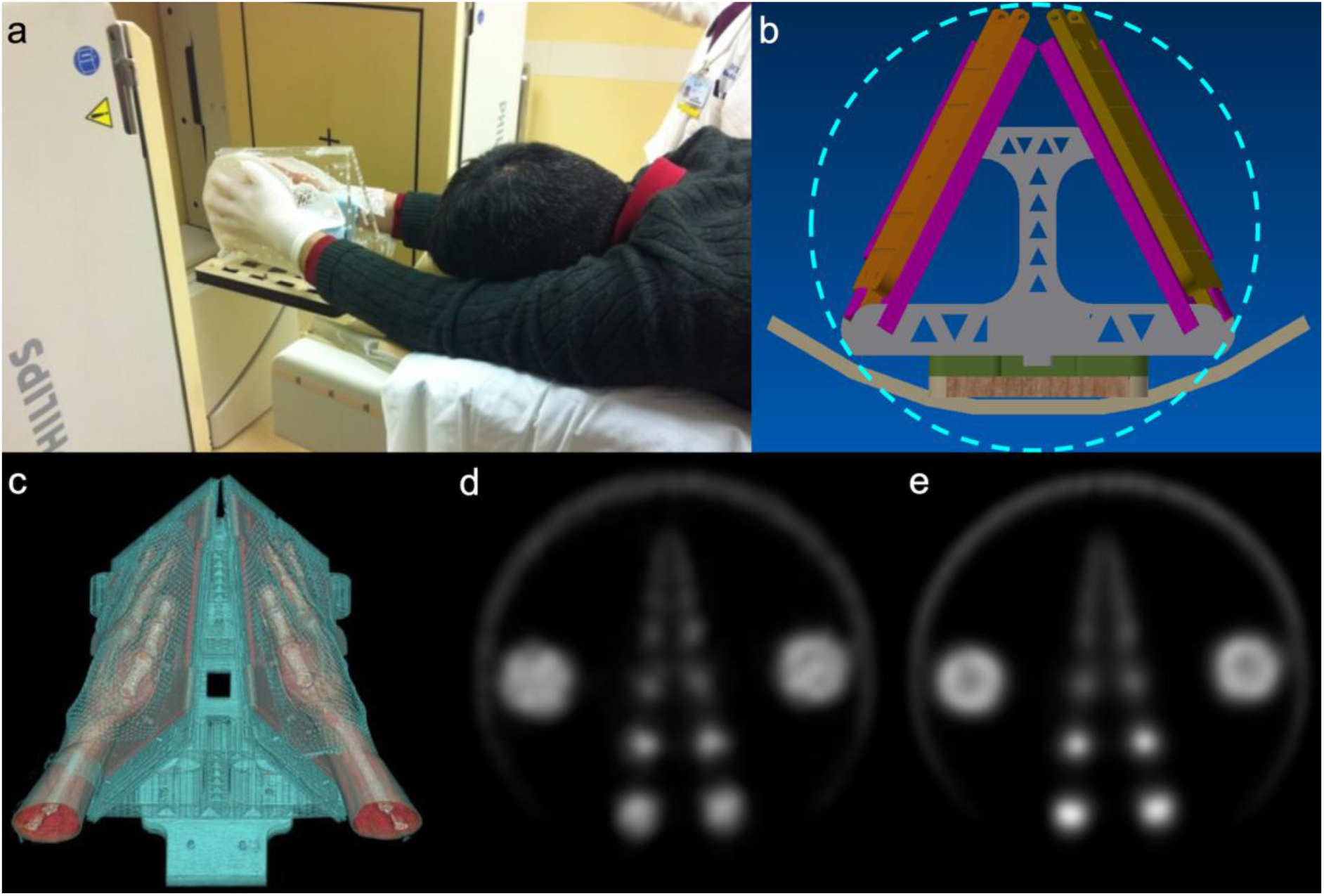
SPECT/CT imaging protocol development. The immobilization device **(a-c)**. Participant position with their hands, inside the immobilizer, outstretched over the shoulders in superman position **(a)**. Design drawing, showing that a circular gamma camera data acquisition trajectory (light blue line) is feasible (b). 3D rendering generated from CT scans of a participant in our pilot study, showing that the base station allows for maintenance of the natural angles of the arms during scanning **(c)**. Comparison of image reconstruction schemes **(d-e)**. OSEM **(d)** and OSEM with PSF correction **(e)**. The largest spheres are of diameter 3.8 cm and show the characteristic ring-like artifact on **(e)**, however, the contrast for smaller spheres (2.2 cm and below) is improved

Next, we evaluated a range of SPECT image reconstruction schemes using a phantom with fillable spheres with varying sizes, that approximately match the sizes of the hand joints, with the goal of obtaining high quantitative reproducibility and contrast-to-noise ratio despite the small size of the joints of the wrist and hand.

Overall our acquisition protocol converged on these acquisition parameters: 64 frames (32 per head), 20 seconds/frame (total ∼11 minutes), zoom 1.46 and matrix size 128 x 128 pixels. With these acquisition parameters, we compared the performance of the iterative ordered subset expectation maximization (OSEM) method and the OSEM method with point spread function (PSF) correction (Fig. 1). We evaluated the reconstructed image properties such as contrast-recovery-coefficient, image uniformity, and the bias-variance trade-off. Accordingly, we selected the OSEM method with PSF correction (4 iterations, 16 subsets), incorporating both attenuation and scatter correction. The reconstructed image slice thickness for SPECT was 3.19 mm while that of CT was 1.0 mm.

### SPECT/CT image acquisition and analysis

All SPECT/CT images were acquired using Philips BrightView XCT (Philips Healthcare, Cleveland, OH) equipped with low-energy all-purpose (LEAP) collimator. Dual-phase scans were acquired starting at 5 min (soft-tissue phase) and 180 min (osseous phase) following a single intravenous injection of 963±36 MBq of 99mTc-MDP. A low-dose CT was acquired for the same field of view after each SPECT scan using an integrated cone-beam x-ray source and a flat-panel detector (tube current: 20 mA; voltage: 120 kVp). The calculated additional radiation dose due to low-dose CT was 0.2 mSv. The total scan time (SPECT+CT) for each phase was 15 min. Patients were allowed to resume their normal activity during the interval between the 5- and 180-min scans.

The reconstructed images were reviewed by one nuclear medicine physician with 15-years of experience on a workstation running OsiriX MD v.8.0 (Pixmeo, Geneva, Switzerland). Twenty joints per hand were assessed (wrist, carpometacarpal [CMC], metacarpophalangeal [MCP], proximal and distal interphalangeal [PIP & DIP]). The wrist joint was considered as a single entity. Images were evaluated qualitatively (visual assessment) and quantitatively. Visual assessment described the signal intensity using a subjective 4-point scale commonly reported in practice (no/mild/moderate/marked uptake). During data analysis, the first two categories were considered within normal and the latter two (moderate or marked uptake) were considered abnormal.

To quantify tracer uptake, a volume of interest (VOI) was drawn on each of the 20 joints based on the low-dose CT images, and the maximum count values were recorded from the corresponding co-registered SPECT images after checking for any mis-registration. Values from all the joints that were both clinically and scintigraphically normal/unremarkable were averaged. The ratio of the maximum count value in each joint normalized by this average, termed maximum corrected count ratio (MCCR), was computed.

For subset analysis, the 11 joints known to be more frequently affected by RA; which included 1^st^-5^th^ MCPs, 2^nd^-4^th^ PIPs, 1^st^ IP and wrist joints; were classified as Group I. The rest of the joints (n=9 per hand), including 1^st^-5^th^ CMC and 2^nd^-4^th^ DIP joints, were classified as Group II.

To ensure consistency and reproducibility, each of the two SPECT phases were reviewed on separate days; also, all the qualitative and quantitative readings were repeated after at least 3-month interval, on the same viewing workstation by the same reader to quantify intra-observer agreement.

### Statistical analysis

Qualitative data were expressed as frequencies and percentages. Association between categorical variables was compared using chi-square test for independent variables or Spearman’s rank correlation, as appropriate. Quantitative data were summarized and expressed as median (range). Two-tailed Mann-Whitney U or Wilcoxon rank test was used for comparing two independent or related groups; respectively. Intra-reader agreement was measured using kappa statistic for qualitative observations and intraclass correlation coefficient (ICC) for quantitative variables. Differences in paired readings was measured using McNemar’s test.

## Results

### Patients

Between May 2015 and May 2016, four participants (all males; median age 67.5 years, range: 53-74) were recruited for this pilot study. Their clinical characteristics are summarized in Table 1. Three participants successfully completed the two scans (at the baseline and 1-month follow-up), while one participant completed only the baseline scan. No patients were excluded due to non-compliance with the protocol. During each visit, SPECT/CT scans during the two phases of radiotracer uptake were successfully completed. No visual evidence of intra-scan motion artifacts was detected in any of the acquired images (4 participants, 7 studies with a total of 14 SPECT/CT scans and 280 joints evaluated).

**Table 1.** Clinical characteristics of the 4 study participants

| Characteristic | Patient 1 | Patient 2 | Patient 3 | Patient 4 |
| --- | --- | --- | --- | --- |
| Age (years) | 74 | 70 | 56 | 65 |
| Antibody status | seropositive | seronegative | seronegative | seronegative |
| Clinically abnormal joints | 7 | 0 | 2 | 2 |
| Baseline DAS-28 | 3.65 | 0.7 | 3.06 | 2.43 |
| Baseline CRP | 14 | 0.7 | 16 | 1.2 |
| Treatments | MTX | SSZ<br>Prednisone<br>Hydroxurea | SSZ | Etanercept |
| DAS-28 at 6 months | 2.33 | 1.11 | 1.99 | 2.43 |
| Final status | Responder | Responder | Non-responder | Non-responder |

*MTX methotrexate, SSZ Sulfasalazine, DAS-28 disease-activity score for 28 joints, CRP C-reactive protein*

### Qualitative and Quantitative SPECT/CT Findings

Among the total 280 joints evaluated, only 22 joints (16 in Group I and 6 in Group II) were clinically abnormal. SPECT imaging showed abnormally increased tracer uptake during both soft-tissue and osseous phases in 10 joints, all from Group I. However, soft-tissue and osseous-phase SPECT showed additional abnormality in 72 (31 Group I, 41 Group II) and 92 (44 Group I, 48 Group II) joints, respectively, that did not present with obvious clinical findings. The pattern of abnormal radiotracer uptake within the same joint was notably different between the soft-tissue and osseous phases (Fig. 2).

**Fig. 2.**
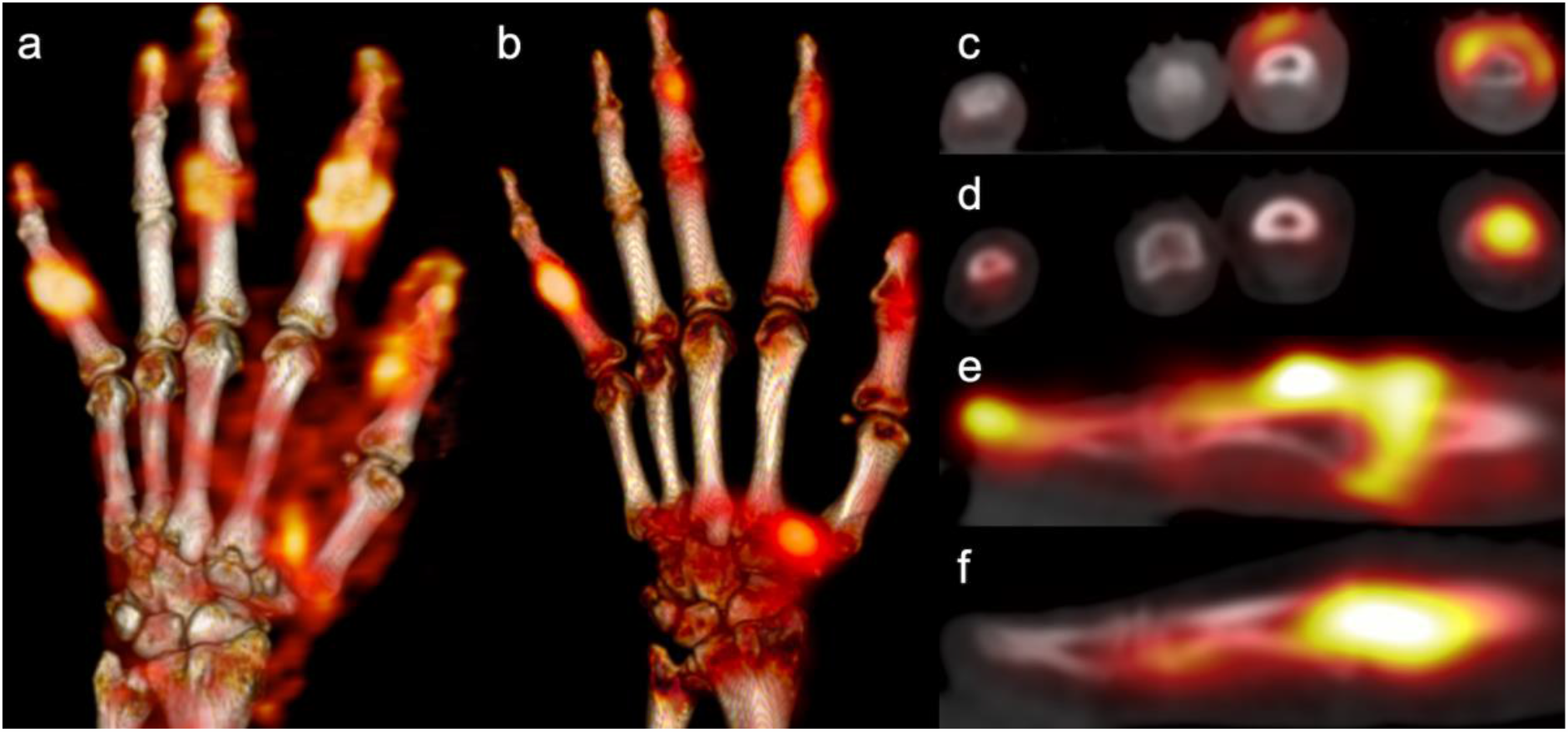
Soft-tissue and osseous SPECT/CT images of RA patient. Volume rendered SPECT/CT images of the left hand of a 74-year-old man with seropositive RA **(a-b)**. Soft-tissue SPECT/CT demonstrating hypervascularity in 1^st^ IP & CMC, 2^nd^, 3^rd^ & 5^th^ PIP joints **(a)**. Osseous phase SPECT/CT showed increased osseous uptake in some of the hypervascular joints (1^st^ CMC, 2^nd^ and 5^th^ PIP; but not the 1^st^ IP and 3^rd^ PIP joints) **(b)**. Axial view of fused SPECT/CT images **(c-d)**. Nearly circumferential hypervascularity pattern around the 2^nd^ PIP and to a lesser extent the 3^rd^ PIP **(c)**. Corresponding image from the osseous SPECT/CT phase demonstrating increased osseous turnover in the 2^nd^ but not the 3^rd^ PIP region **(d)**. Sagittal view of magnified fused SPECT/CT images of the 2^nd^ PIP joint **(e-f)**. Linear soft tissue hypervascularity along the extensor aspect of the joint **(e)**. Corresponding osseous phase image demonstrating asymmetric tracer distribution across the joint, being more prominent on the distal end of the proximal phalanx of the index finger **(f)**. The corresponding CT images (not shown) showed no erosive involvement

Quantitatively, the mean MCCR from soft-tissue phase SPECT was significantly higher for clinically abnormal joints (2.50±1.68) compared to clinically normal ones (1.35±0.79; *P* < 0.0001). Such differences were not seen with the osseous phase SPECT assessments, which showed a median MCCR of 1.76±1.21 and 1.34±0.71 for clinically abnormal and normal joints, respectively.

MCCR demonstrated significant positive correlation with the 4-point ordinal visual score of uptake intensity, with Spearman’s rank correlation coefficients of 0.78 (95 CI%: 0.73-0.82) and 0.84 (95% CI: 0.8-0.87) for the soft-tissue and osseous-phase SPECTs, respectively. Qualitative and quantitative SPECT findings are summarized in Table 2.

**Table 2.**
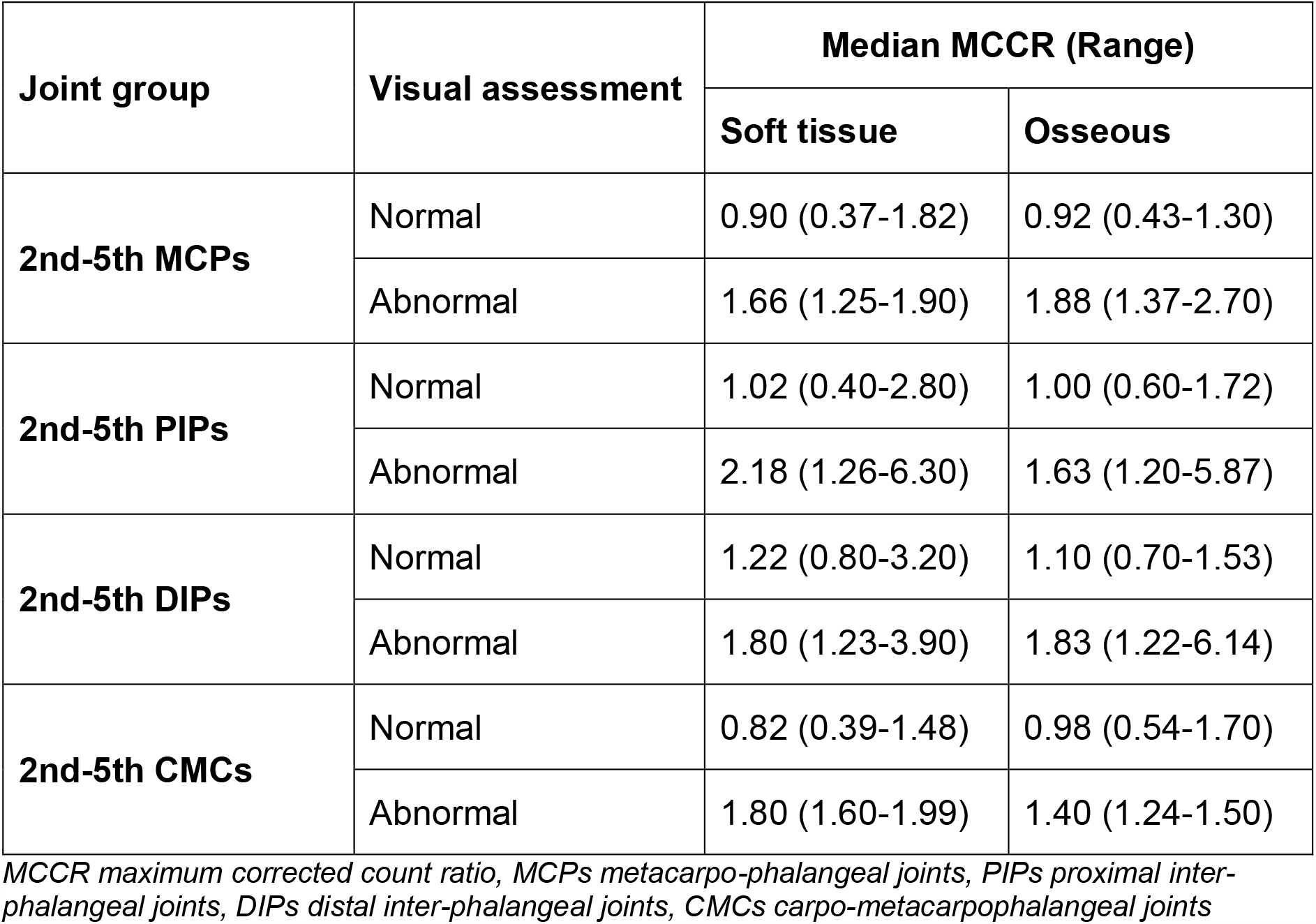
Qualitative and quantitative SPECT findings in different joint groups

### Agreement between soft tissue and osseous phases

Soft-tissue MCCR showed moderate positive correlation with osseous-phase MCCR (r = 0.64, 95%CI = 0.56-0.70; *P* < 0.001). Visually, the presence of soft tissue hypervascularity was not accompanied with osseous uptake in 17/82 joints (8 from Group I, 9 from Group II). Conversely, 37/102 joints with high osseous uptake did not demonstrate hypervascularity (21 from Group I, 16 from Group II). That discordance between the soft-tissue and osseous phases was more evident within Group I (*P* = 0.03) compared to Group II joints (*P* = 0.2).

### Observer agreement

MCCR was highly reliable (ICC > 0.995) when a repeated measurement was performed (Table 3). Intra-observer agreement on the qualitative SPECT findings was substantial for all joints, both from soft-tissue and osseous phases (Table 3).

**Table 3.** Intra-observer agreement on qualitative and quantitative evaluations of the different joints from dual phase 99mTc-MDP SPECT/CT of the hands

| Joints | N | Soft-tissue phase |  | Osseous phase |  |
| --- | --- | --- | --- | --- | --- |
|  |  | Kappa<br>(95% CI) | ICC<br>95% CI | Kappa<br>95% CI | ICC<br>95% CI |
| All | 280 | 0.777<br>(0.723-0.830) | 0.979<br>(0.973-0.983) | 0.700<br>(0.642-0.759) | 0.979<br>(0.973-0.983) |
| Group I | 154 | 0.799<br>(0.729-0.868) | 0.980<br>(0.973-0.985) | 0.762<br>(0.690-0.835) | 0.993<br>(0.990-0.995) |
| 2nd-5th<br>PIPs | 56 | 0.803<br>(0.694-0.912) | 0.979<br>(0.964-0.988) | 0.720<br>(0.603-0.837) | 0.994<br>(0.990-0.997) |
| 2nd-5th<br>MCPs | 56 | 0.799<br>(0.652-0.945) | 0.966<br>(0.943-0.980) | 0.795<br>(0.668-0.922) | 0.988<br>(0.980-0.993) |
| Group II | 126 | 0.747<br>(0.664-0.830) | 0.977<br>(0.967-0.984) | 0.626<br>(0.532-0.719) | 0.993<br>(0.991-0.995) |
| 2nd-5th<br>DIPs | 56 | 0.665<br>(0.516-0.814) | 0.970<br>(0.949-0.982) | 0.568<br>(0.440-0.696) | 0.992<br>(0.986-0.995) |
| 2nd-5th<br>CMCs | 56 | 0.629<br>(0.431-0.828) | 0.960<br>(0.932-0.976) | 0.423<br>(0.231-0.616) | 0.976<br>(0.959-0.986) |
| 1st CMCs | 14 | 0.779<br>(0.556-1.000) | 0.958<br>(0.876-0.987) | 0.853<br>(0.651-1.000) | 0.996<br>(0.987-0.999) |

*MCPs* metacarpo-phalangeal joints, *PIPs* proximal inter-phalangeal joints, *DIPs* distal inter-phalangeal joints, *CMCs* carpo-metacarpophalangeal joints

### Relation of quantitative SPECT to response

Group I joints showed higher MCCR from baseline soft tissue SPECT scans (Table 4) in non-responders compared to responders (1.65 vs. 1.31).

**Table 4.**
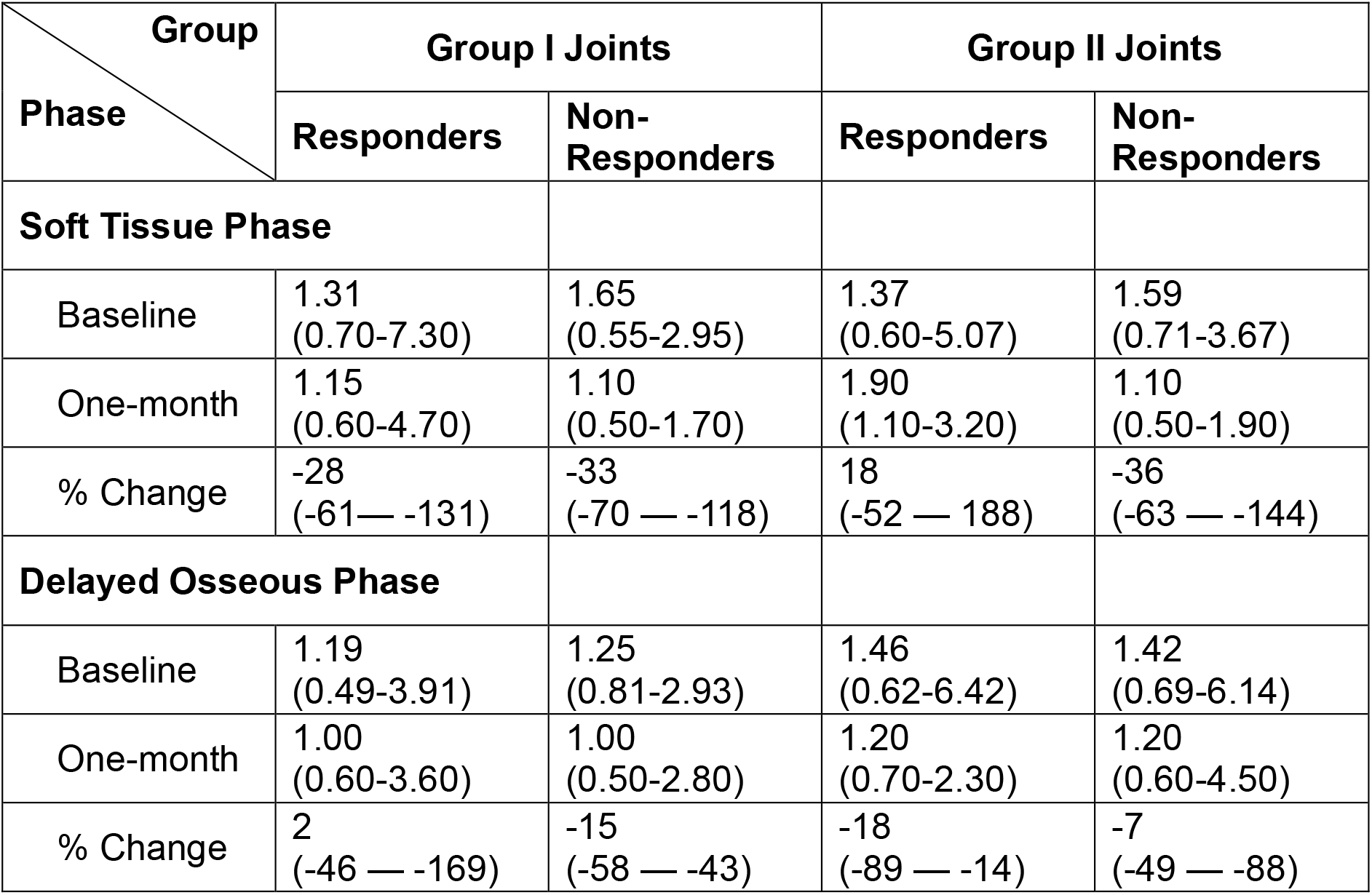
Maximum corrected count ratio (MCCR) at the baseline and 1-month after therapy for group I (rheumatoid arthritis) and Group II (osteoarthritis) joints in responder and non-responder patients

Similarly, a slightly higher MCCR from baseline delayed osseous phase SPECT scans was noted in non-responders compared to responders (1.25 vs. 1.19; Table 4).

## Discussion

In RA a quantitative understanding of synovial vascularity and altered bone metabolism may provide means for robustly assessing disease activity before irreversible anatomical damage is manifested (6, 19). In this prospective pilot study, we demonstrated the feasibility and reliability of measuring markers corresponding to synovitis and bone metabolism in small joints of an RA cohort using dual-phase SPECT/CT scanning. Our rationale is that SPECT/CT scans, due to their inherent 3D nature, eliminate the superimposition problem of planar imaging, and therefore improve the sensitivity, specificity and spatial resolution compared to planar scintigraphy (20).

Previous studies designed a specialized multi-pinhole high resolution collimator for use in animal models (14) and RA patients (15, 16, 21). Multipinhole cameras are built on conventional SPECT machines and provide significantly higher sensitivity and resolution compared to parallel-hole collimators, but typically have a vastly limited field of view (10 or 20 cm depends on aperture design), which limits the scans to the fingers or wrist of one hand at a time. SPECT scans using this approach were able to detect a larger number of diseased joints compared to planar imaging. Also, investigators accurately localized the exact sub-region of the joint that showed increased uptake. However, studies performed using such cameras to date employed only the single delayed osseous phase SPECT scanning for the most clinically affected hand. That may represent a particular limitation for RA assessment, as RA is typically bilateral and often symmetric disease. Also, software fusion with other cross-sectional images is challenging.

Hybrid SPECT/CT machines have the advantages of hardware fusion of the functional tracer distribution and anatomical information in three dimensions, which allows accurate registration, attenuation correction and potential for assessment of CT changes within the small joints (22). The added radiation dose from CT was not significant (∼0.2 mSv).

Our results demonstrated that, quantitatively, the tracer activity from soft-tissue uptake (indicative of vascularity) was significantly higher in clinically-positive joints. On the other hand, osseous uptake was noted in 44 RA joints that were clinically unremarkable, 24 of which showed corresponding hypervascularity in soft-tissue phase. The sensitivity and specificity of clinical signs and DAS-28 have been a concern for not being able to differentiate disease activity from chronic inactive inflammation. In comparison to tools that measure vascularization like doppler ultrasound, subclinical disease is easily missed during clinical evaluation (23-25). Although the high sensitivity of this SPECT/CT method may help evaluate the disease burden, the findings should be interpreted in the clinical context (26), with emphasis on the pattern and distribution of the tracer (15).

Because soft tissue and osseous SPECT/CT phases reflect different pathophysiologic processes, it is not surprising to encounter discordance between the two scans. Overall, 41 joints in Group I demonstrated hypervascular soft-tissue uptake, of them 8 did not show any abnormality on the osseous phase. Isolated blood pool soft tissue hyperemia has been described for detecting RA-synovitis (13, 27). On the other hand, 21/54 joints with increased osseous metabolism did not demonstrate corresponding hypervascularity. Chronic inactive arthritis can demonstrate persistent osseous uptake which limit the specificity of this finding (12, 28). However, it is worth mentioning that these isolated osseous changes could be the result of some other pathogenic processes in the context of RA disease. For example, early bony alteration have been demonstrated using diphosphonate and were not depicted on MRI (8, 12, 15, 29) which may reflect reactive bone repair for pre-erosive lesions. In previous studies, the joints that became eroded within 2 years were scintigraphically active and showed no radiographic evidence of erosive changes at the baseline (8); while a negative bone scan was prognostic for the absence of inflammatory joint disease for up to 3.6 years (30). Alternatively, active osteitis in RA patients has been postulated to start preferentially from the bone marrow rather than the synovial membrane (inside-out theory) (5).

In this pilot work, we demonstrated the feasibility of extracting reproducible qualitative and quantitative measures of the soft tissue vascularity and osseous metabolism of both hands’ joints in a single scan. This information may be currently not available by other modalities. The ultimate aim of our current work would therefore be to develop sensitive biomarkers for detecting the earliest reversible pathophysiologic changes that could benefit from a specific line of treatment, like tumor necrosis factor alpha (TNF-α) blockers (31).

Our study has some merits, which include a prospective design, innovative approach for hand positioning, which allowed both hands to be imaged simultaneously, utilizing both soft-tissue and osseous-phase SPECT, and detailed analysis of observer agreement. There were also limitations. First, there was no independent validation of the results using other imaging modalities or biopsy. Second, given the pilot nature of this study, our sample size was small. Finally, our analysis considered each joint or joint category as independent entity, and did not explicitly account for correlation between joints in the same participant.

In conclusion extracting reliable quantitative and qualitative measures from dual-phase 99mTc-MDP SPECT/CT of the hands is feasible in patients with RA. These measures could unleash important pathologic information on soft-tissue vascularity and bone metabolism, both of high relevance to RA assessment before and after therapy.

## Data Availability

Data can be made available on a reasonable request to the corresponding author

## Acknowledgements

The authors would like to acknowledge the contributions of Drs. Piotr Maniawski from Philips Healthcare, John Brock from the University of California Davis and Dr. Angela Da Silva for helpful discussions regarding the content of the manuscript.

